# Associations of hearing loss with social isolation, loneliness, and depressive symptoms among older adults in the Health, Aging, and Body Composition Study

**DOI:** 10.64898/2026.08.06.26359904

**Authors:** Mary C. Thoma, Erin Ferguson, Jacqueline M. Torres, Kristine Yaffe, Nicole M. Armstrong, Jennifer A. Deal, Danielle Powell, Bonnielin K. Swenor, Willa D. Brenowitz

## Abstract

**Background:** Hearing loss (HL) may be a risk factor for poor psychosocial outcomes among older adults, but evidence remains mixed. We assessed associations of self-reported and objective HL with and without hearing aid use with social contact, loneliness, and depression pooled across 6 years of follow-up.

**Methods:** We studied 2049 Black and White adults from the Health, Aging, and Body Composition study aged 70-79 at recruitment. Self-reported HL and audiometric HL with and without hearing aid use were assessed at analytic baseline (Year 5, 2001-2002). Outcomes were frequency of contact with family and friends (<weekly vs. at least weekly), depressive symptoms (CESD-10), and loneliness (CESD-10 item “I felt lonely”) measured across 6 annual visits. Adjusted for demographic and clinical variables, we used generalized linear regression with generalized estimating equations to assess associations with outcomes pooled across six follow-up waves.

**Results:** Self-reported HL (16%) was associated with more depressive symptoms (ß=0.13 SD; 95%CI:0.03,0.24), but no other outcome. Objective HL without hearing aid use (11%) was associated with infrequent contact with friends (OR=1.38; 95%CI:1.07,1.78) and more depressive symptoms (ß=0.19 SD; 95%CI:0.07,0.31); objective HL with hearing aid use (9%) was not associated with these outcomes. Objective HL, regardless of hearing aid use, was borderline associated with more frequent feelings of loneliness.

**Discussion:** Objective HL without hearing aid use may be an important risk factor for isolation from friendship networks and depressive symptoms among older adults. Self-reported HL and objective HL with hearing aid use may also be linked to some adverse psychosocial outcomes.

## INTRODUCTION

Hearing loss is common among older adults, affecting more than 60% of adults over the age of 70 in the United States (1,2). Age-related hearing loss is characterized by difficulty understanding speech through noise, such as in crowded rooms (3–5). Social isolation, a growing global epidemic with a multitude of health consequences, is also common in older adults. It has been associated with increased mortality, risk of cardiovascular disease, cognitive decline, and psychosocial outcomes such as loneliness and depression (6–9). Adults with age-related hearing loss may start to avoid social situations as communication becomes difficult, leading to social isolation (10). Hearing loss may also increase loneliness and depression via adverse impacts on the quantity or quality of social interactions (11).

While prior research has found associations of hearing loss with isolation measures (12–15) and psychosocial outcomes such as depressive symptoms (14,16,17) and loneliness (16,18–20), not all have. Studies have been varied in their designs and measurements. One important source of variability is how hearing loss itself is defined and measured. Many studies rely on self-reported measures of hearing loss (12–14,17,18,21–23), while few studies have used objective measures such as audiometric assessments (15). Self-reported measures capture the subjective experience of hearing loss. Objective measures, such as audiometric assessments, capture the level of audiometric hearing loss, which may or may not be apparent to the person. Studies comparing the prevalence of self-reported hearing loss vs. audiometrically assessed hearing loss have shown that older adults are likely to underestimate their hearing loss (24,25). Meanwhile, adults who overestimate their hearing loss are more likely to experience anxiety and depression (24). Studies comparing the relationships between self-reported versus objective hearing and psychosocial outcomes are limited (16,22). Comparing the associations of self-reported vs. objective hearing loss with psychosocial outcomes may be important for understanding the full impact of hearing loss on social and emotional well-being among older adults, as the associations may differ depending on whether hearing loss is subjectively (self-reported) versus objectively (audiometrically) assessed.

Furthermore, there is a limited body of research assessing the association of hearing aid use and social isolation, loneliness, or depression. A recent review of studies found mixed results in changes in social isolation and loneliness after hearing aid interventions, though results generally showed improvement in isolation and loneliness (26). The authors of this review noted a lack of quality evidence due lack of control for confounders, short follow-up periods, and small sample sizes (26). Few longer-term observational studies of hearing loss and social isolation, loneliness, or depressive symptoms have investigated associations with hearing aid use (16,27).

We assessed longitudinal associations of hearing loss with frequency of contact with friends, frequency of contact with family, loneliness, and depressive symptoms in a community-based study of Black and White older adult from the Health, Aging, and Body Composition Study (Health ABC). We evaluated differences between objective and subjective hearing measures, as well as hearing loss with and without hearing aid use. Because hearing loss is more prevalent in White men than other groups (1,28) and some studies show differences by sex (12,15,16,18), we also assessed potential differences by race and sex. We hypothesized that self-reported hearing loss, which may better reflect one’s overall experience of hearing loss than audiometric assessments, would have stronger associations with social contact, loneliness, and depression than objective hearing loss. We additionally hypothesized that hearing aid use would be associated with better outcomes than no hearing aid use among participants with hearing loss.

## METHODS

### Study population

The Health ABC Study is a longitudinal community-based cohort of 3075 Black and White adults age 70-79 with no reported difficulty in walking ¼ mile or climbing one flight of stairs at recruitment. Participants were recruited between 1997-1998 from select zip codes in Pittsburgh, Pennsylvania and Memphis, Tennessee. Participants were followed with annual clinical visits and semiannual telephone interviews to update health and functional status (29). Self-reported and objective hearing assessments were first conducted in study Year 5 (completed between 2001-2002), which served as our analytic baseline. To be eligible for inclusion in our analysis, participants had to have self-reported and objective hearing assessments at analytic baseline (Year 5) and at least one measure of frequency of social contact, loneliness, or depressive symptoms at any of the follow-up visits (study Years 6 through 11). Of those who were initially recruited, 342 participants did not participate in Year 5 data collection, 560 participants were excluded due to incomplete hearing data, 98 participants had no available outcome data, and 26 participants were excluded due to incomplete covariate data, for a total analytic sample of 2049. Differences in characteristics between participants who were included compared to those excluded are shown in Supplemental Table 1.

### Hearing loss

Objective hearing loss was measured using pure tone audiometry tested in a sound-treated booth. Audiometric thresholds were taken at 0.5, 1, 2, and 4 kHz. Hearing loss was defined as a pure tone average (PTA) of >40 decibels in the better hearing ear across these thresholds, following other studies, which corresponds to moderate to severe hearing loss (28,30). Audiometric assessments were completed without use of hearing aids. Separately, participants self-reported use of hearing aids in daily life. Objective hearing loss was categorized into three groups: (1) hearing loss with reported use of hearing aids, (2) hearing loss without reported use of hearing aids, and (2) no hearing loss, which served as the reference group.

Participants additionally self-reported whether they could easily converse in a crowded room with a hearing aid if necessary and whether they felt that any difficulty with their hearing limited their personal or social life. For consistency with prior studies, inability to converse in a crowd was used as a measure of self-reported hearing loss (12–14). For exploratory purposes, we additionally assessed the association of perceived limitations due to hearing difficulties with our outcomes of interest. All measures of hearing were assessed at Year 5.

### Psychosocial outcomes

Participants were asked how often in a typical week they get together with children and relatives as well as with friends and neighbors. Possible answers ranged from “at least once per day” to “less than once per week”. We created separate measures of infrequent social contact with family and infrequent social contact with friends, each defined as getting together less than once per week vs. once a week or more. Frequency of contact was measured at study Years 6 and 8 through 11.

Depressive symptoms were measured on a continuous scale using the Center of Epidemiologic Studies 10-item Depression Scale (CESD-10) (31). Loneliness was measured using the CESD-10 item, “I felt lonely”. Participants reported how frequently they felt lonely in the past week from 1 (“rarely or none of the time”) to 4 (“most or all of the time”). Depressive symptoms and loneliness were z-scored and modeled as continuous outcomes. Loneliness and depression were measured at Years 6, 8, 10, 11.

### Covariates

Demographic covariates included sex (male/female), race (Black/White), and education (less than high school/high school degree) as recorded at Year 1 as well as age at analytic baseline (Year 5). We also controlled for participants’ history of clinical conditions cumulative up to Year 5, including heart disease, stroke, diabetes, and hypertension. These conditions were assessed via a combination of participant interview, medical records, medications, and baseline laboratory values. In models of loneliness and depressive symptoms, we controlled for repeated measures of contact with family and friends in sensitivity analyses to evaluate whether associations were independent of social contact.

### Statistical Analyses

We modeled the associations of self-reported and objective hearing loss with subsequent infrequent contact with family, infrequent contact with friends, loneliness, and depressive symptoms. We pooled outcome measures across the six annual visits following the analytic baseline and used generalized estimating equations to account for the non-independence of repeated outcome assessments. We specified general linear regression models with a logit link for binary outcomes (infrequent contact with family, friends) and an identity link for continuous outcomes (depressive symptoms, frequency of loneliness), an exchangeable working correlation structure, and robust standard errors (32). Models were adjusted for all baseline covariates.

We additionally ran each model with an interaction term between self-reported or objective measures of hearing loss and sex or race, as well as models stratified by sex or race. To assess whether the associations between hearing loss and our outcomes of interest varied over time, we additionally ran each model with an interaction term between hearing loss measures and time. Time was continuous from Year 5 to Year 11.

Finally, we assessed the associations of reported hearing aid use (yes/no) with our outcomes of interest among participants with objective hearing loss.

## RESULTS

At analytic baseline, participants were an average of 77.4 years of age (SD: 2.8). One fifth (21%) of participants had objective hearing loss at analytic baseline; 16% self-reported hearing loss. Among participants with objective hearing loss, 45% reported use of hearing aids. Regardless of hearing aid use, those with objective hearing loss were more likely to be older, male, and have a history of diabetes than those without hearing loss. Those with objective hearing loss who used hearing aids were more likely to be White, have at least a high-school education, and have a history of stroke compared to both those with unaided hearing loss and no hearing loss. Those with hearing loss but no hearing aids were less likely to have a high-school education and less likely to have a history of stroke than those with no hearing loss (Table 1).

**Table 1.** Analytic baseline (Year 5) characteristics of Health ABC participants included in at least one model.

| **Baseline characteristics** | **No HL**<br>**(N=1627)** | **HL with HA use**<br>**(N=191)** | **HL with no HA use**<br>**(N=231)** |
| --- | --- | --- | --- |
| Age, years (SD) | 77.20 (2.76) | 78.62 (2.90) | 78.07 (3.01) |
| Female | 923 (56.7%) | 68 (35.6%) | 85 (36.8%) |
| Black | 642 (39.5%) | 29 (15.2%) | 71 (30.7%) |
| High school degree | 1296 (79.7%) | 160 (83.8%) | 146 (63.2%) |
| History of heart disease | 460 (28.3%) | 59 (30.9%) | 68 (29.4%) |
| History of stroke | 139 (8.5%) | 18 (9.4%) | 15 (6.5%) |
| History of diabetes | 382 (23.5%) | 54 (28.3%) | 60 (26.0%) |
| History of hypertension | 1409 (86.6%) | 163 (85.3%) | 196 (84.8%) |
| Self-reported HL | 208 (12.8%) | 58 (30.4%) | 64 (27.7%) |
| Self-reported limitations due to HL | 128 (7.9%) | 65 (34.4%) | 58 (25.1%) |
| Pure-tone average (SD) | 24.96 (8.50) | 52.89 (9.04) | 48.65 (9.04) |

Note. HL=Hearing loss. HA=Hearing aid.

Among participants with objective hearing loss, those who reported use of hearing aids had worse objective hearing loss, i.e. higher PTA, than those who reported no use of hearing aids (PTA= 52.9, SD=9.0 vs. PTA=48.7, SD=9.0) (Table 1). They also were more likely to self-report hearing loss, i.e. report difficulty conversing in a crowded room even when using hearing aids, and to report limitations in their personal or social life due to hearing difficulty, compared to those who reported no use of hearing aids (self-reported hearing loss: 30.4% vs. 27.7%, respectively; perceived limitations due to hearing loss: 34.4% vs. 25.1%, respectively) (Table 1, Figure 1). Among those with self-reported hearing loss, 63% had no objective hearing loss, 18% had objective hearing loss with reported hearing aid use, and 19% had objective hearing loss with no reported hearing aid use (Figure 1).

**Figure 1.**
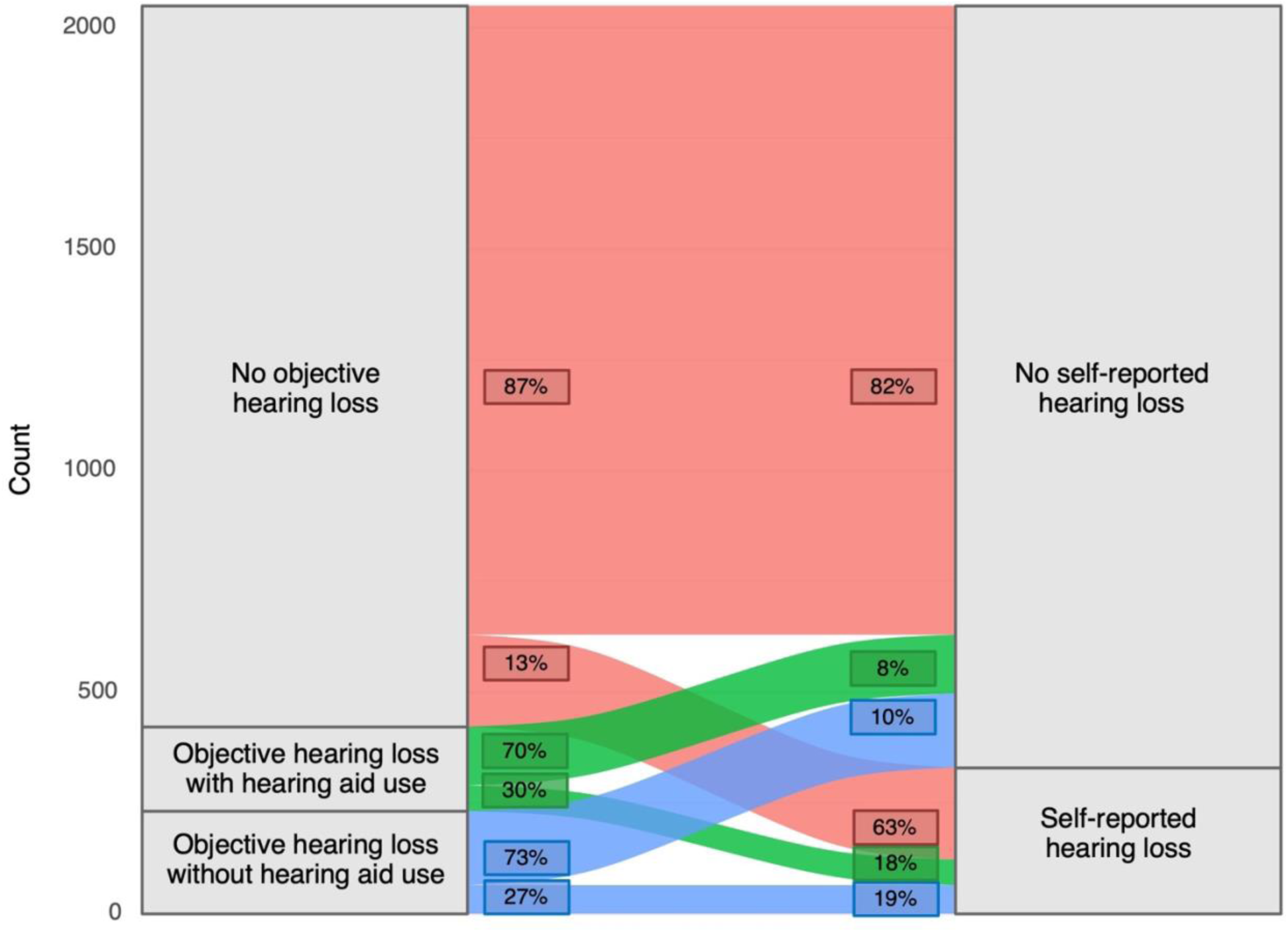
Overlap of objective and self-reported hearing loss among Health ABC participants at study year 5 (2001-2002), N=2049.

### Hearing loss and infrequent contact with friends and family

Compared to no objective hearing loss, objective hearing loss without hearing aid use was associated with greater odds of having infrequent contact with friends (OR=1.38, 95% CI: 1.07-1.78) but was not associated with contact with family (OR=0.91, 95% CI: 0.71-1.16); objective hearing loss *with* hearing aid use was not associated with either outcome (Figure 2). Compared to no self-reported hearing loss, self-reported hearing loss was not associated with frequency of contact with friends but was borderline associated with *lower* odds of low family contact (OR=0.85, 95% CI: 0.69, 1.04) (Figure 2). Self-reported limitations in one’s personal or social life due to hearing difficulties were not associated with contact with family or friends, compared to reporting no limitations (Figure 2).

**Figure 2.**
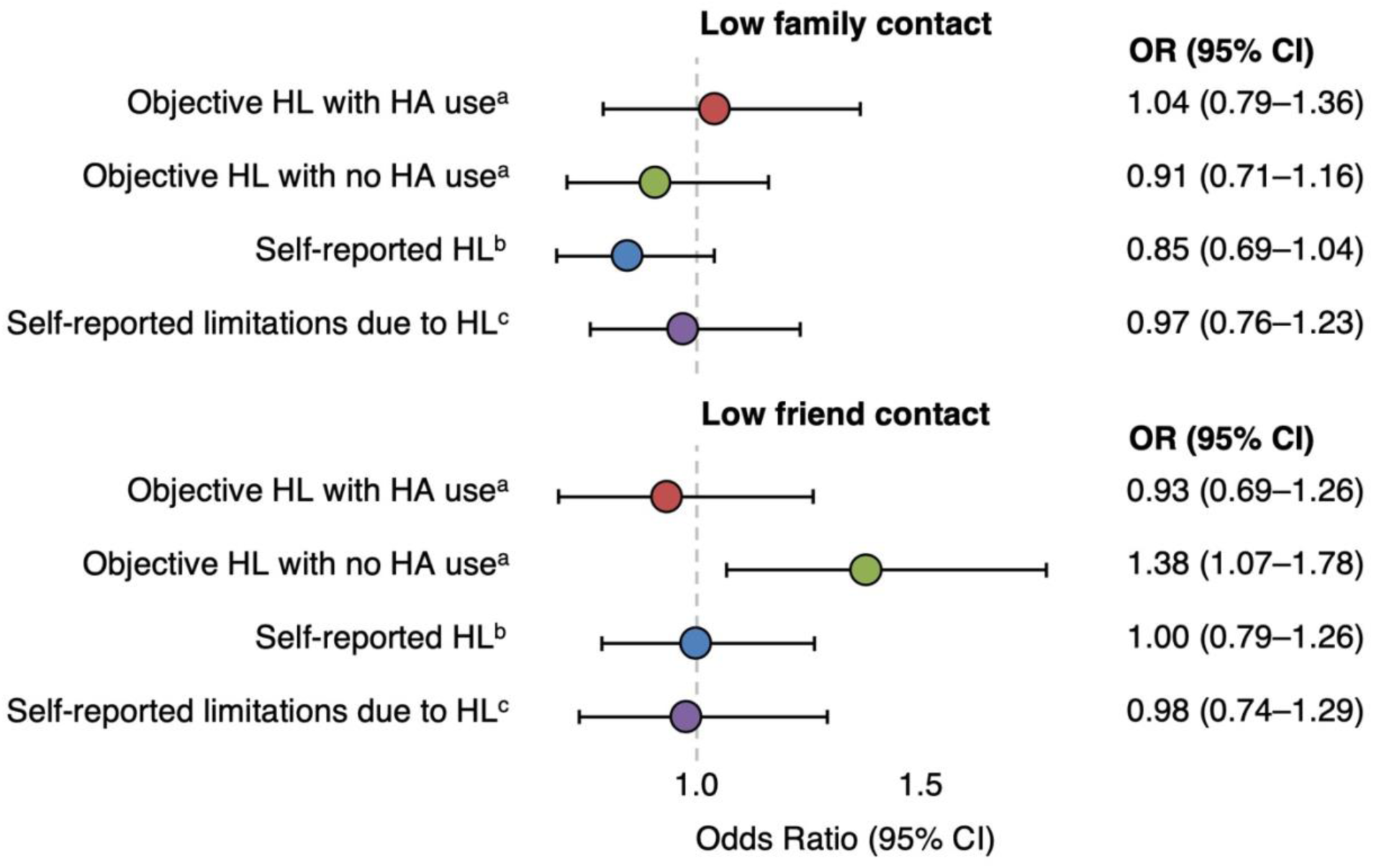
Associations of objective and self-reported hearing loss with odds of low frequency of contact (<weekly) with family and with friends among Health ABC participants. HL=hearing loss, HA=hearing aid. ^a^Reference group = no objective hearing loss, N in model=2018 ^b^Reference group = no self-reported hearing loss, N in model=2018 ^c^Reference group = no self-reported limitations, N in model=2015

### Hearing loss and depressive symptoms

Compared to no objective hearing loss, objective hearing loss without hearing aid use was associated with more depressive symptoms (ß=0.19, 95% CI: 0.06, 0.32), while objective hearing loss *with* hearing aid use was not (Figure 3). Self-reported hearing loss (compared to no self-reported hearing loss) and self-reported limitations in one’s personal and social life due to hearing difficulties (compared to no limitations) were also each associated with more depressive symptoms (self-reported hearing loss: ß=0.13, 95% CI: 0.03, 0.24; perceived limitations due to hearing loss: ß=0.34 SD, 95% CI: 0.22, 0.46) (Figure 3). Results were similar when adjusting for time-varying contact with family and friends (S. Table 1).

**Figure 3.**
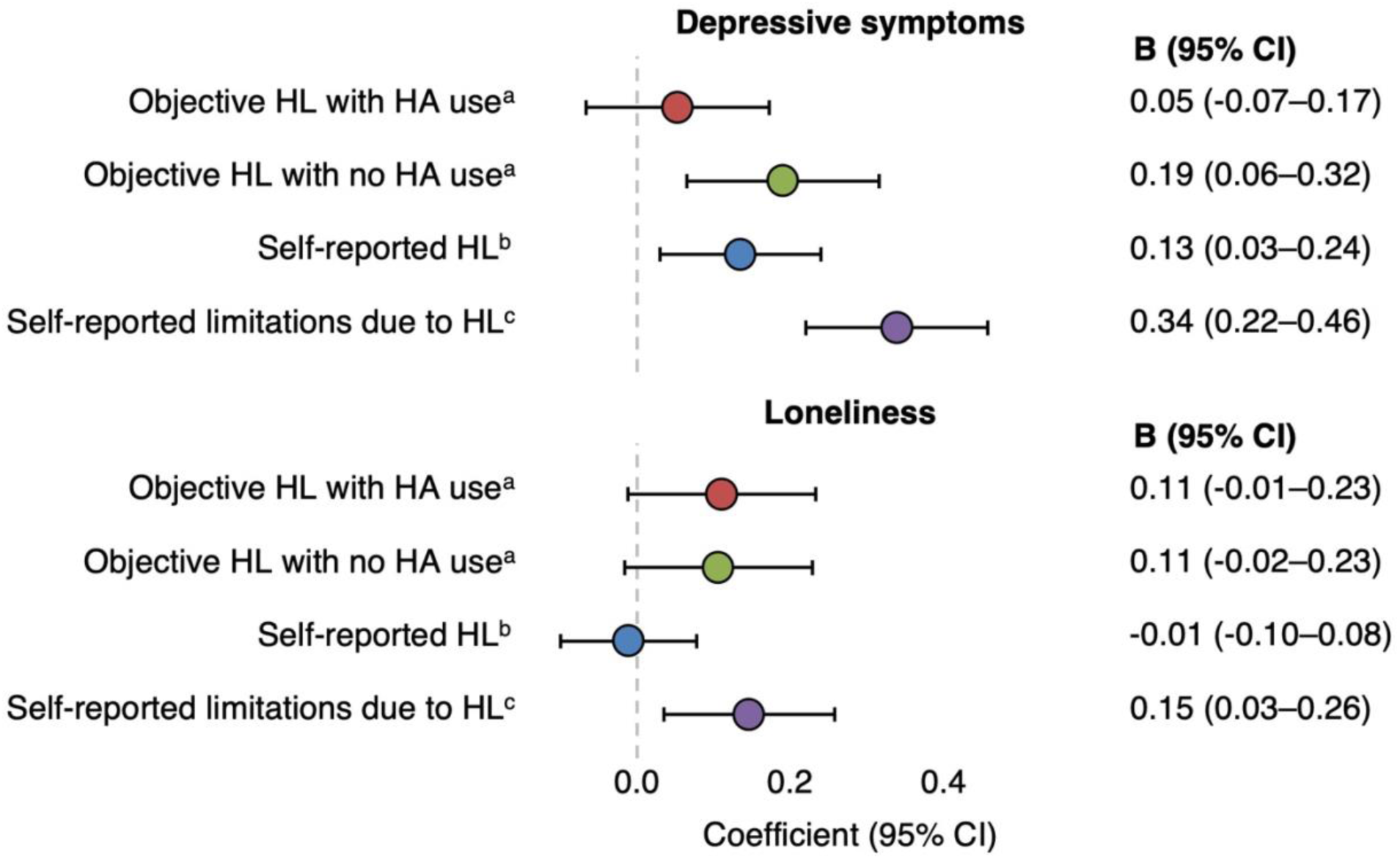
Associations of self-reported hearing limitations and objective aided and unaided hearing impairment with depressive symptoms and loneliness (z-scored) among Health ABC participants. HL=hearing loss, HA=hearing aid. ^a^Reference group = no objective hearing loss, N in model=2049 ^b^Reference group = no self-reported hearing loss, N in model=2049 ^c^Reference group = no self-reported limitations, N=2046

### Hearing loss and loneliness

Compared to no objective hearing loss, hearing loss, both with and without hearing aid use, was borderline associated with a greater frequency of loneliness (hearing loss *with* hearing aid use: ß=0.11, 95% CI: −0.01, 0.23; hearing loss *without* hearing aid use: ß=0.11, 95% CI: −0.02, 0.23) (Figure 3). Self-reported hearing loss was not associated with feelings of loneliness (Figure 3). Participants who reported that hearing difficulties led to limitations in their personal and social life had more frequent feelings of loneliness (ß=0.15 SD, 95% CI: 0.03, 0.26) compared to participants who reported no limitations (Figure 3). In general, estimates were slightly attenuated but similar when controlling for time-varying frequency of contact with family and friends (S. Table 2). However, the association between hearing loss without hearing aid use and loneliness was much closer to the null when controlling for time-varying frequency of contact with family and friends (ß=0.06, 95% CI: −0.07, 0.19) (S. Table 2).

**Table 2.** Prevalence of outcomes measured at Year 6 by exposure status among Health ABC participants included in at least one model.

|  | N Missing | Objective HL |  |  | Self-reported HL |  |
| --- | --- | --- | --- | --- | --- | --- |
|  |  | No HL | HL with HA use | HL with no HA use | No | Yes |
| Low family contact | 154 | 503 (33.4%) | 62 (34.4%) | 66 (31.4%) | 537 (33.9%) | 94 (30.1%) |
| Low friend contact | 153 | 229 (15.2%) | 30 (16.7%) | 35 (16.7%) | 247 (15.6%) | 47 (15.0%) |
| Loneliness score (SD) | 26 | 1.34 (0.66) | 1.39 (0.70) | 1.36 (0.68) | 1.34 (0.66) | 1.36 (0.69) |
| Depressive symptoms (SD) | 28 | 4.91 (4.26) | 5.11 (4.47) | 5.27 (4.67) | 4.86 (4.25) | 5.52 (4.70) |

Note. Low family/friend contact = less than once per week. Range of loneliness scores = 1-4. Range of depression scores = 0-30. HL=Hearing loss. HA=Hearing aid.

### Interactions of hearing loss with time on the associations of frequency of contact, depression, loneliness

There was not a significant interaction with time for most outcomes (S. Table 3). There was a significant interaction between self-reported hearing loss and continuous time since analytic baseline in the association with frequency of loneliness (ß of interaction term= −0.02 SD, 95% CI: −0.04, −0.002). Self-reported hearing loss was not associated with loneliness at Year 6 (one year after self-reported hearing was measured) (ß=0.02, 95% CI: −0.08, 0.11) or Year 11 (six years after self-reported hearing was measured) (ß= −0.10, 95% CI: −0.21, 0.02), although trends were towards an inverse association.

### Interactions of hearing loss with sex and race on the associations with frequency of contact, depression, loneliness

Interaction terms between hearing loss and sex and race were not significant. However, in models stratified by sex, objective hearing loss without aid use (compared to no hearing loss) was associated with greater odds of infrequent contact with friends among women (OR=1.57, 95% CI: 1.05, 2.35), while the magnitude of association was smaller and not significant among men (OR=1.27, 95% CI: 0.91, 1.77). Likewise, the association between objective hearing loss without hearing aid use and depressive symptoms was stronger among women (ß=0.27, 95% CI: 0.05, 0.49) than among men (ß=0.14, 95% CI: −0.01, 0.28). Self-reported hearing loss was associated with lower odds of infrequent contact with family among Black participants (OR=0.61, 95% CI: 0.40, 0.91) but not among White participants (OR=0.95, 95% CI: 0.74, 1.21). Similarly, Black participants with objective hearing loss with and without hearing aid use had lower odds of infrequent contact with family compared to those with no objective hearing loss, although these associations were imprecise and not statistically significant (hearing loss *with* hearing aid use: OR=0.50, 95% CI: 0.20, 1.22; hearing loss *without* hearing aid use: OR=0.85, 95% CI: 0.53, 1.35). Among White participants, objective hearing loss *with* hearing aid use was associated with more frequent feelings of loneliness (ß=0.16, 95% CI: 0.02, 0.29) but no association was observed among Black participants (ß= −0.04, 95% CI: −0.28, 0.21).

### Associations of hearing aid use with frequency of contact, depression, loneliness, among participants with objective hearing loss

In sensitivity analyses restricted to participants with objective hearing loss (max N=422), hearing aid use was not significantly associated with any outcome. However, all associations were in the expected direction, aligning with results from main models in which no hearing loss was the reference. For example, hearing aid use was associated with lower odds of infrequent contact with friends (OR=0.75, 95% CI: 0.52, 1.08) and fewer depressive symptoms (ß= −0.12, 95% CI: −0.28, 0.04). No association was observed between hearing aid use and contact with family (OR=1.13, 95% CI: 0.80, 1.60) or frequency of loneliness (ß=0.00, 95% CI: −0.16, 0.17).

## DISCUSSION

We assessed the associations of hearing loss and hearing aid use with contact with friends and family and with loneliness and depressive symptoms across 6 years of follow up in a cohort of community-dwelling Black and White older adults in two U.S. metropolitan regions. Compared to no objective hearing loss, objective (i.e., audiometrically assessed) hearing loss without hearing aid use was associated with greater odds of infrequent contact with friends and more depressive symptoms across follow-up. In contrast, objective hearing loss *with* hearing aid use was not associated with these outcomes. Objective hearing loss, regardless of hearing aid use, was borderline associated with more frequent feelings of loneliness, although these associations were not statistically significant. Self-reported hearing loss (i.e., ability to converse in a crowded room using hearing aids if necessary) was not associated with contact with children or family or with feelings of loneliness but was associated with more depressive symptoms, compared to no self-reported hearing loss. Associations with hearing loss did not change substantially over time for most outcomes, indicating persistent associations with psychosocial outcomes.

Our findings extend upon and are consistent with prior studies noting associations with hearing loss and psychosocial measures (12–15,17,21,23). We expand upon the literature by directly comparing associations of objective versus subjective hearing loss measurements and hearing aid use with psychosocial outcomes. Interestingly, we found differences in the prevalence of self-reported versus objective hearing loss, as well as in their associations with social contact, loneliness, and depressive symptoms. Consistent with prior findings, many older adults self-reported hearing loss despite having no measurable audiometric hearing loss, while others with audiometric hearing loss, both with and without hearing aid use, reported no hearing loss (24,25). We expected that self-reported hearing loss would have greater associations with isolation, loneliness, and depression, as we theorized that participants who were aware of their hearing loss were more likely to withdraw from social scenarios. Surprisingly, objective hearing loss with no hearing aid use was associated with greater odds of infrequent contact with friends, whereas self-reported hearing loss was not. Both self-reported and objective hearing loss without hearing aid use were associated with more depressive symptoms.

Interestingly, perceived personal and social limitations due to hearing loss were not associated with frequency of contact with family or friends, but were associated with greater depressive symptoms and loneliness. It is possible that older adults who perceive limitations due to hearing loss do not alter their social habits but experience lower quality interactions and thus feel lonelier and have more depressive symptoms. It is also possible that we are not capturing subtle differences in social contact between groups, such as time spent together per outing, or size of social network. Our results suggest that hearing loss may have an impact on frequency of contact with friends whether or not the hearing loss is perceived, and that perceived limitations due to hearing loss have the strongest association with depressive symptoms and feelings of loneliness.

Our findings add to mixed evidence on the associations between hearing aid use and psychosocial outcomes. Our results support prior evidence indicating that associations between objective hearing loss without hearing aid use is associated with adverse social and depressive outcomes compared to those with no hearing loss, but that older adults with hearing loss who use hearing aids have similar outcomes as those with no hearing loss (17,33). Recent trial evidence demonstrated a reduction in social isolation and loneliness after a hearing aid intervention among older adults with hearing loss (34); our work extends such findings to a longer follow-up duration. In sensitivity analyses restricted to participants with objective hearing loss, associations between hearing aid use and psychosocial outcomes were not significant, possibly due to very small sample size. However, the direction and magnitude of associations are suggestive of the protective role of hearing aid use. For example, hearing aid use was associated with lower odds of infrequent contact with friends and fewer depressive symptoms.

Contrary to recent trial evidence (34) but in-line with prior literature (35,36), we did not find an association between hearing aid use and loneliness among participants with objective hearing loss. Compared to those with no hearing loss, both those with objective hearing loss *with* hearing aid use and those with objective hearing loss *without* hearing aid use had similar risk of greater frequency of loneliness. Together, our findings suggest that treatment of hearing loss with hearing aids may reduce social isolation and depressive symptoms but may have little impact on loneliness. Future studies should assess these associations in larger sample sizes.

Finally, we note mixed evidence that associations varied by sex and race. Objective hearing loss without hearing aid use was associated with low friend contact and more depressive symptoms in women; the magnitude of these associations were smaller and not significant among men. These findings align with results from prior studies (12,15,16,18). To our knowledge, prior studies have not assessed differences in association by race. We found that among White participants, objective hearing loss without hearing aid use was associated with more frequent feelings of loneliness, but no association was observed among Black participants. Interestingly, among Black participants self-reported hearing loss was associated with lower odds of infrequent contact with family. These findings are exploratory due to the small sample sizes and suggest inconsistent differences in associations between hearing loss and psychosocial outcomes within different demographic groups, but should be studied in larger samples.

We recognize several important limitations. Hearing was only assessed at a single time point, limiting our ability to capture incident or progressive hearing loss over time. Our measures of social contact only capture daily to less than weekly visits with family or friends; this may not capture meaningful variation in social contact or identify more socially isolated individuals. Our assessment of loneliness was drawn from a single item on the Center for Epidemiologic Studies Depression (CES-D) Scale, which, while commonly used, is not as comprehensive as other validated measures of loneliness (37,38). Finally, Health ABC enrolls only Black and White participants and is a relatively healthy cohort. Our analytic sample size was made up of a higher proportion of White, well-educated, and healthy participants than the original Health ABC sample, partly due to death and drop out before hearing was audiometrically assessed at Year 5. The generalizability of our findings may therefore be limited.

Our study benefits from several important strengths. We incorporated both objective and subjective assessments of hearing loss, allowing us to compare the strength of associations between perceived hearing loss and actual hearing loss with social isolation and psychological outcomes. Furthermore, we compare use of hearing aids in a biracial and well-characterized cohort of older adults. Finally, while most prior studies have relied on cross-sectional data, we use longitudinal data to demonstrate that the observed associations persist across follow-up. Our results highlight the long-term psychosocial impact of hearing loss among older adults and suggests potential benefits of hearing aid use.

## Acknowledgements

We thank Health ABC participants and staff who made this research possible. Author contributions – Conceptualization: all authors; Data curation: Mary C. Thoma, Kristine Yaffe, Willa D. Brenowitz; Formal analysis: Mary C. Thoma, Erin Ferguson, Willa Brenowitz; Funding acquisition: Jacqueline M. Torres, Willa D. Brenowitz; Methodology/interpretation: all authors; Validation: Erin Ferguson; Visualization: Mary C. Thoma, Willa D. Brenowitz; Writing (original draft): Mary C. Thoma, Willa D. Brenowitz; Writing (review & editing): all authors

## Data availability

Health ABC data is available for approved research proposals (see https://healthabc.nia.nih.gov/)

