## Supplementary material for "Associations of hearing loss with social isolation, loneliness, and depressive symptoms among older adults in the Health, Aging, and Body Composition Study"

### Contents:

**Supplemental Table 1.** Characteristics at study baseline (Year 1) of Health ABC participants.

**Supplemental Table 2.** Associations of hearing limitations and hearing impairment with depression and loneliness, controlling for time-varying contact with family and friends.

**Supplemental Table 3.** Interactions between time and associations of hearing limitations and hearing impairment.

**Supplemental Table 4.** Associations of hearing aid use with frequency of contact with outcomes of interest among older adults with audiometric hearing loss (pure tone average>40).

**Supplemental Table 1.** Characteristics at study baseline (Year 1) of Health ABC participants.

| Year 1 characteristics | Included in analytic sample (N=2049) | Excluded from analytic sample (N=1026) |
|---|---|---|
| Age, years (SD) | 73.43 (2.83) | 74.02 (2.93) |
| Female | 1076 (52.5%) | 508 (49.5%) |
| Black | 742 (36.2%) | 539 (52.5%) |
| High school degree | 1602 (78.2%) | 690 (67.8%) |
| History of heart disease | 479 (23.4%) | 343 (33.4%) |
| History of stroke | 148 (7.2%) | 99 (10.0%) |
| History of diabetes | 1011 (49.3%) | 580 (56.5%) |
| History of hypertension | 1263 (61.6%) | 713 (69.5%) |
| Self-reported limitations due to HL | 180 (8.8%) | 89 (8.8%) |
| Low family contact | 644 (31.9%) | 276 (27.7%) |
| Low friend contact | 368 (18.2%) | 227 (22.8%) |
| Loneliness (SD) | 1.22 (0.56) | 1.26 (0.59) |
| Depressive symptoms (SD) | 2.90 (3.29) | 3.44 (3.57) |

HL=Hearing loss

**Supplemental Table 2.** Associations of hearing limitations and hearing impairment with depression and loneliness, controlling for time-varying contact with family and friends among older adults in the Health ABC study. N in models=1961.

| | Depressive symptoms (SD) | | | | Loneliness (SD) | | | |
|---|---|---|---|---|---|---|---|---|
| | B | LCL | UCL | P-val | B | LCL | UCL | P-val |
| Objective hearing loss (ref= none) | | | | | | | | |
| With reported hearing aid use | 0.06 | -0.07 | 0.20 | 0.35 | 0.10 | -0.04 | 0.23 | 0.15 |
| Without reported hearing aid use | 0.14 | 0.01 | 0.28 | 0.04 | 0.06 | -0.07 | 0.19 | 0.34 |
| Self-reported hearing loss | 0.13 | 0.02 | 0.24 | 0.03 | -0.01 | -0.11 | 0.09 | 0.84 |
| Perceived limiations due to hearing loss | 0.35 | 0.22 | 0.47 | 0.02 | 0.14 | 0.02 | 0.26 | 0.02 |

**Supplemental Table 3.** Interactions between time and associations of hearing limitations and hearing impairment with frequency of contact with family and friends (N in models=2018) and with depressive symptoms and loneliness (N in models=2049) among older adults in the Health ABC study.

| | Low family contact | | | | Low friend contact | | | | Depressive symptoms (SD) | | | | Loneliness (SD) | | | |
|---|---|---|---|---|---|---|---|---|---|---|---|---|---|---|---|---|
| | OR | LCL | UCL | P-val | OR | LCL | UCL | P-val | B | LCL | UCL | P-val | B | LCL | UCL | P-val |
| Self-reported hearing loss * Time | 1.01 | 0.94 | 1.08 | 0.81 | 1.07 | 0.97 | 1.17 | 0.16 | 0.00 | -0.02 | 0.016 | 0.71 | -0.02 | -0.04 | -0.002 | 0.03 |
| Objective hearing loss (ref= none) * Time | | | | | | | | | | | | | | | | |
| With reported hearing aid use | 1.04 | 0.96 | 1.12 | 0.37 | 0.95 | 0.83 | 1.09 | 0.46 | -0.01 | -0.04 | 0.01 | 0.25 | 0.02 | -0.01 | 0.05 | 0.17 |
| Without reported hearing aid use | 1.00 | 0.91 | 1.09 | 0.92 | 1.07 | 0.97 | 1.19 | 0.19 | 0.01 | -0.02 | 0.03 | 0.73 | 0.01 | -0.02 | 0.04 | 0.61 |

**Supplemental Table 4.** Associations of hearing aid use with frequency of contact with family and friends (N in models=417) and with depressive symptoms and loneliness (N in models=422) among older adults with audiometric hearing loss (pure tone average>40) in the Health ABC study.

| | Low family contact | | | | Low friend contact | | | | Depressive symptoms (SD) | | | | Loneliness (SD) | | | |
|---|---|---|---|---|---|---|---|---|---|---|---|---|---|---|---|---|
| | OR | LCL | UCL | P-val | OR | LCL | UCL | P-val | B | LCL | UCL | P-val | B | LCL | UCL | P-val |
| Reported hearing aid use | 1.13 | 0.80 | 1.60 | 0.48 | 0.75 | 0.52 | 1.08 | 0.13 | -0.12 | -0.28 | 0.042 | 0.15 | 0.00 | -0.16 | 0.17 | 0.97 |
